# The use of Artificial Intelligence in the out of hospital care settings: A Scoping Review

**DOI:** 10.1101/2025.04.04.25325245

**Authors:** Jamie Miles, Mike Brady, Leanne Smith, Charlotte Cotterill, Charlotte Levey

## Abstract

**Background:** Out of hospital services face significant challenges, including growing patient demand, workforce limitations, and evolving care pathways. Artificial Intelligence (AI) technologies offer potential solutions, but their application in out-of-hospital settings remains inconsistently implemented and poorly understood.

**Objectives:** To identify the types of AI technologies being applied in out-of-hospital settings, explore their purposes and implementation contexts, and examine associated outcomes.

**Methods:** Six electronic databases were searched for English-language studies published between 2013-2024. Eligible studies involved AI technologies in the out-of-hospital emergency services setting. Data were synthesised according to five implementation domains: system level, dispatch zone, response zone, on-scene zone, and onward prognosis.

**Results:** From 236 publications, we identified diverse AI applications across the care pathway. System-level implementations (46 studies) featured AI for demand forecasting, optimal resource allocation, and strategic facility location, with demonstrated improvements in coverage efficiency of 10-20%. In the dispatch zone (32 studies), AI-enhanced emergency call triage and ambulance allocation reduced response times by up to 10-20%. Response-level applications (43 studies) included intelligent traffic management and real-time route optimisation, reducing travel times by 15-30%. On-scene zone implementations (75 studies) supported clinical decision-making with cardiac arrest rhythm detection, achieving an area under the curve (AUC) values exceeding 0.90 and acute coronary syndrome prediction sensitivities of 85-90%. Onward prognosis models (19 studies) predicted patient outcomes with AUC values of 0.80-0.90 for survival forecasting, enabling better resource allocation and early intervention. Further inferential analysis applications (21 studies) were also identified that provide higher-level insights through secondary analyses of out-of-hospital data.

**Conclusions:** AI demonstrates significant potential across the care pathway, from operational optimisation to clinical decision support. Future development should focus on real-time adaptive systems, ethical implementation, improved data integration across the care continuum, and rigorous evaluation of real-time patient outcomes. Cross-disciplinary collaboration and standardised reporting of AI implementations will be essential to realise the full potential of these technologies in improving out-of-hospital care delivery.

**Key Messages:** *What is already known on this topic:*

- AI technologies show theoretical promise for enhancing out-of-hospital care services, but limited information exists about their implementation and real-world impact.

*What this study adds:*

- This first comprehensive scoping review identifies a critical implementation gap: of 236 publications describing AI applications in out-of-hospital care, fewer than 15% report functional clinical deployments, and fewer than 5% document sustained implementation with evaluation.

*How this study might affect research, practice or policy:*

- Our findings suggest an urgent need to shift focus from developing novel AI applications to implementing and evaluating existing ones, addressing key barriers, including technical integration challenges, regulatory hurdles, evidence requirements, and organisational change management.

## Introduction

### Rationale

Out of hospital care encompasses diverse services ranging from ambulance (emergency medical services) to NHS 111 providers and out-of-hours care. This system handles over 67 million patient interactions annually in the United Kingdom (UK) alone [1] and faces mounting pressures from an ageing population, stretched workforces, limited community alternatives, growing elective waiting lists, and underutilisation of emerging technologies [2, 3].

The traditional model of ambulances transporting patients to hospitals has undergone a paradigm shift, revealing whole-system challenges requiring comprehensive responses [4]. Artificial Intelligence (AI) represent promising approaches to monitoring, simulating, predicting, and optimising system performance for improved patient outcomes.

### Objectives

The development and implementation of AI methods in this setting remains uneven and slower than anticipated. Multiple barriers exist, including ethical concerns, technological limitations, liability issues, regulatory constraints, and data readiness challenges [5].

Additionally, the lack of interoperable systems within and between services means that transport to emergency departments often remains the path of least resistance despite recognised inefficiencies.

Current ambitions for AI include enhanced triage processes, predictive analytics, telemedicine applications, and remote patient monitoring [6]. However, significant knowledge gaps persist regarding the impact of AI in healthcare [7], necessitating an improved understanding of where these tools are being used, how they are implemented, and their effects on stakeholders [8].

This scoping review aims to address these knowledge gaps by (1) identifying the types of AI technologies being applied in out-of-hospital settings, (2) exploring the purposes for which these technologies are utilised, and (3) examining the outcomes associated with their implementation. The findings will highlight areas for future research specifically relating to urgent and emergency care pathways.

## Methods

### Protocol and registration

The research team developed and approved a protocol but did not formally register it. The initial draft was generated using AI tools [9] and subsequently refined through researcher input, balancing computational efficiency with human insight [10].

### Eligibility criteria

Studies were included if they: (1) related to out-of-hospital emergency care settings (e.g., ambulance services; (2) utilised, piloted, or explored AI technologies; (3) were published in English between 2013-2024; and (4) appeared in peer-reviewed journals, conference proceedings, or reputable preprints. Studies not meeting these criteria were excluded.

### Information sources and search

Six electronic databases were searched: CINAHL, MEDLINE (via PubMed), IEEE Xplore, ACM Digital Library, Scopus, and arXiv. The search strategy combined terms related to out-of-hospital care and AI/machine learning. The full search strategy can be found in the supplementary material.

### Selection of sources of evidence

Records were imported into Rayyan software [11] for screening and de-duplication. Titles and abstracts were independently screened by at least two reviewers, followed by full-text screening for eligible studies. Disagreements were resolved through discussion and consensus.

### Data items

Data extracted included study details (author, year, country), AI technology used, the setting type, primary aim and methodology, outcomes assessed, and key findings/conclusions. Two reviewers performed Data extraction independently, with discrepancies resolved through team discussion.

### Critical appraisal of individual sources of evidence

A formal critical appraisal was not conducted as this is a scoping review. However, study quality and relevance were assessed based on study design, completeness of reporting, and potential biases. Studies with incomplete data or unclear methodologies were flagged for cautious interpretation.

### Synthesis of results

Data were synthesised using a narrative approach, mapping AI applications to their intended functions within out-of-hospital care and were categorised into five domains: system level, dispatch zone, response zone, on-scene zone, and onward prognosis. Quantitative and qualitative findings were summarised according to these domains, effectiveness, and implementation barriers.

## Results

### Selection of sources of evidence

The search yielded a total of 517 records. After removing duplicates, 382 unique studies were screened at the title and abstract levels. Following a full-text review, 236 studies met the inclusion criteria and were included in the final synthesis. The study selection followed the PRISMA-ScR flow diagram methodology (Figure 1).

**Figure 1.**
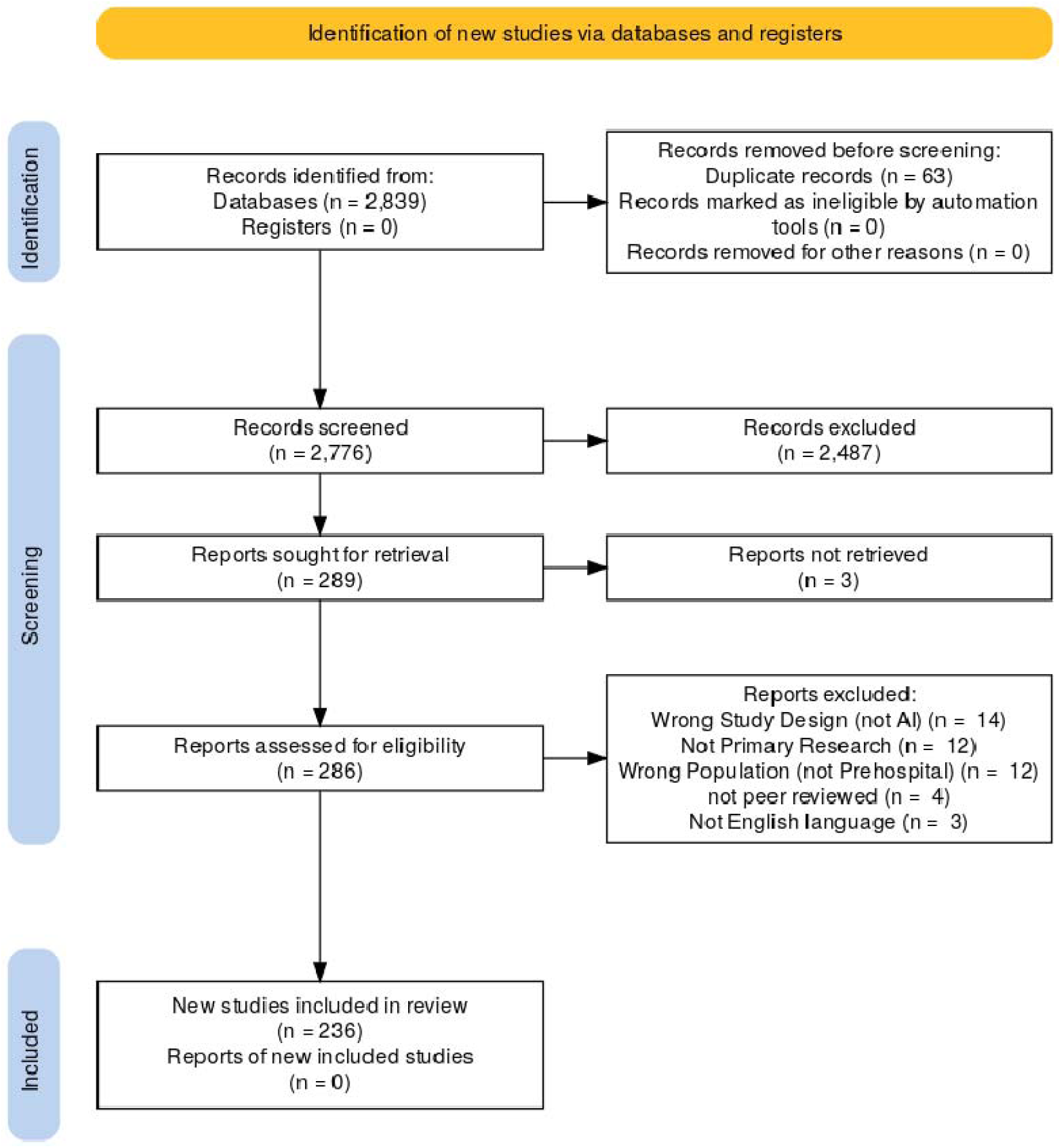

### Characteristics of sources of evidence

The 236 publications originated from 41 countries, with Asia (41%), Europe (28%), and North America (19%) being the dominant regions, led by the United States, Japan, and India (Figure 2). Publication volume increased substantially after 2018, but notably, no significant multi-country collaborations were identified, suggesting limited international knowledge sharing in this field.

**Figure 2.**
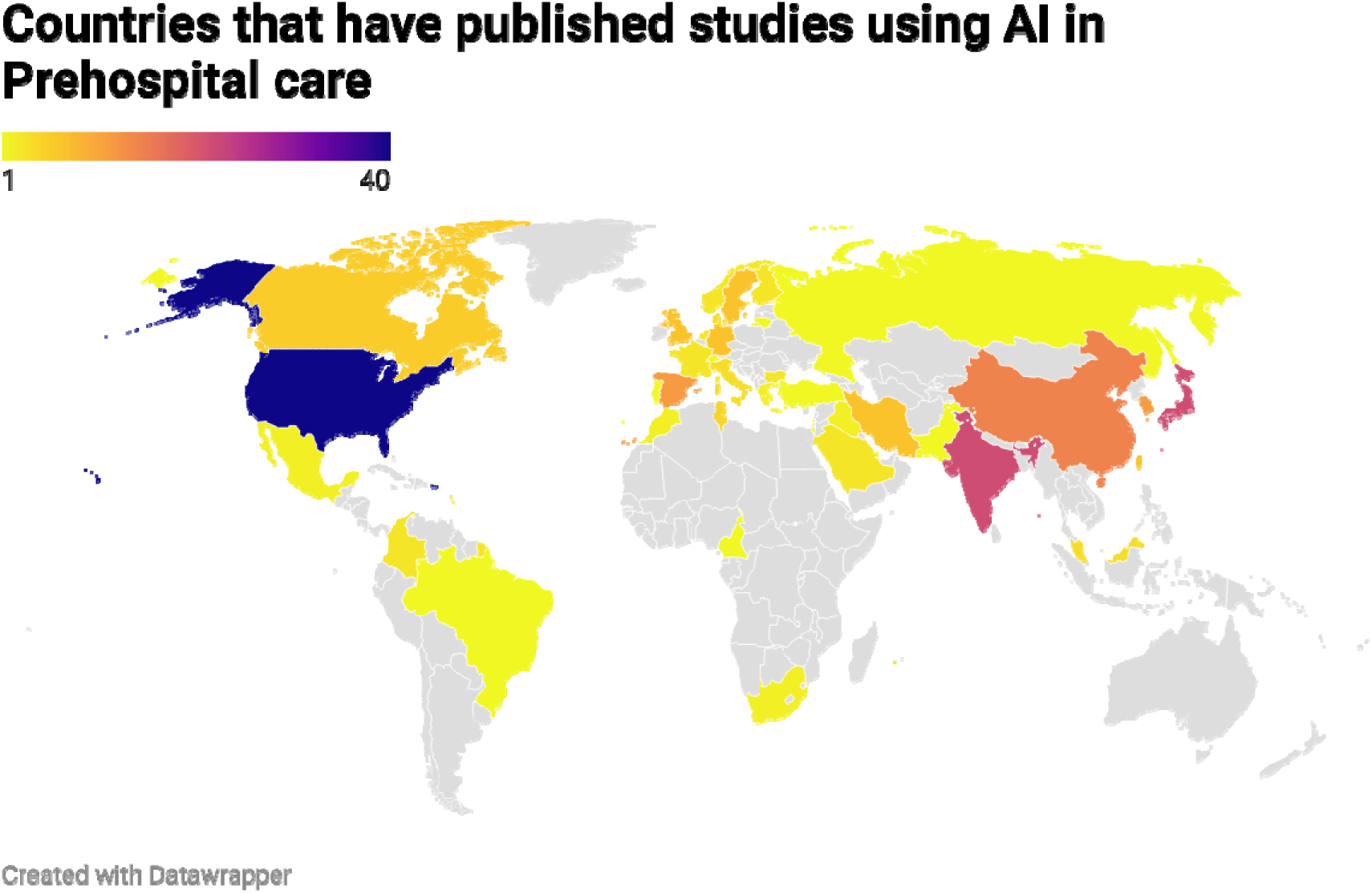

### Network analysis

Network analysis of author collaborations revealed a substantial but relatively sparse collaboration structure within AI out-of-hospital research (1,218 unique authors; 4,799 collaboration links; network density = 0.006475). The field is characterised by several prominent research clusters, with most studies (98.7%) involving multi-author teams (mean = 5.2 authors per study). The analysis identified four key researchers based on publication frequency: Irusta (10 papers), Aramendi (9), Ong (7), and Rawshani (6). The strongest collaborative relationship was between Aramendi and Irusta (9 co-authored papers), while Ong demonstrated the highest degree of centrality (54 unique collaborators), suggesting a role as a significant bridge between research communities.

The network’s structural characteristics—multiple distinct clusters with limited inter-cluster collaboration—suggest opportunities for increased cross-team knowledge exchange to advance the field. This collaboration pattern aligns with the interdisciplinary nature of AI out-of-hospital research, which requires technical, clinical, and methodological expertise integration.

### Synthesis of results

In out-of-hospital care, a generally linear pathway was identified that begins when someone accesses help and ends when they receive definitive care. Our analysis revealed five distinct domains where AI is being applied:

1. System Level (46 studies): Applications optimising the entire infrastructure, including demand forecasting, resource planning, and strategic facility location.
2. Dispatch Zone (32 studies): Tools for initial triage, resource decision-making, and allocation during the call-handling phase.
3. Response Zone (43 studies): Technologies supporting vehicle routing, traffic management, and alternative response modes during transit to patients.
4. On-Scene Zone (75 studies): Applications supporting clinical assessment, intervention, and patient decision-making.
5. Onward Prognosis (19 studies): Models predicting patient outcomes to inform care decisions and resource allocation.

Additionally, we identified Inferential Analysis applications (21 studies) that provide higher-level insights through secondary analyses of out-of-hospital data, examining trends, associations, and underlying mechanisms across the pathway.

Figure 3 illustrates the relationship between these domains across the out-of-hospital care pathway. The following sections detail each domain’s AI applications, outcomes, and challenges. Table 1 shows a matrix of counts between the applied area and the methodological group.

**Figure 3.**
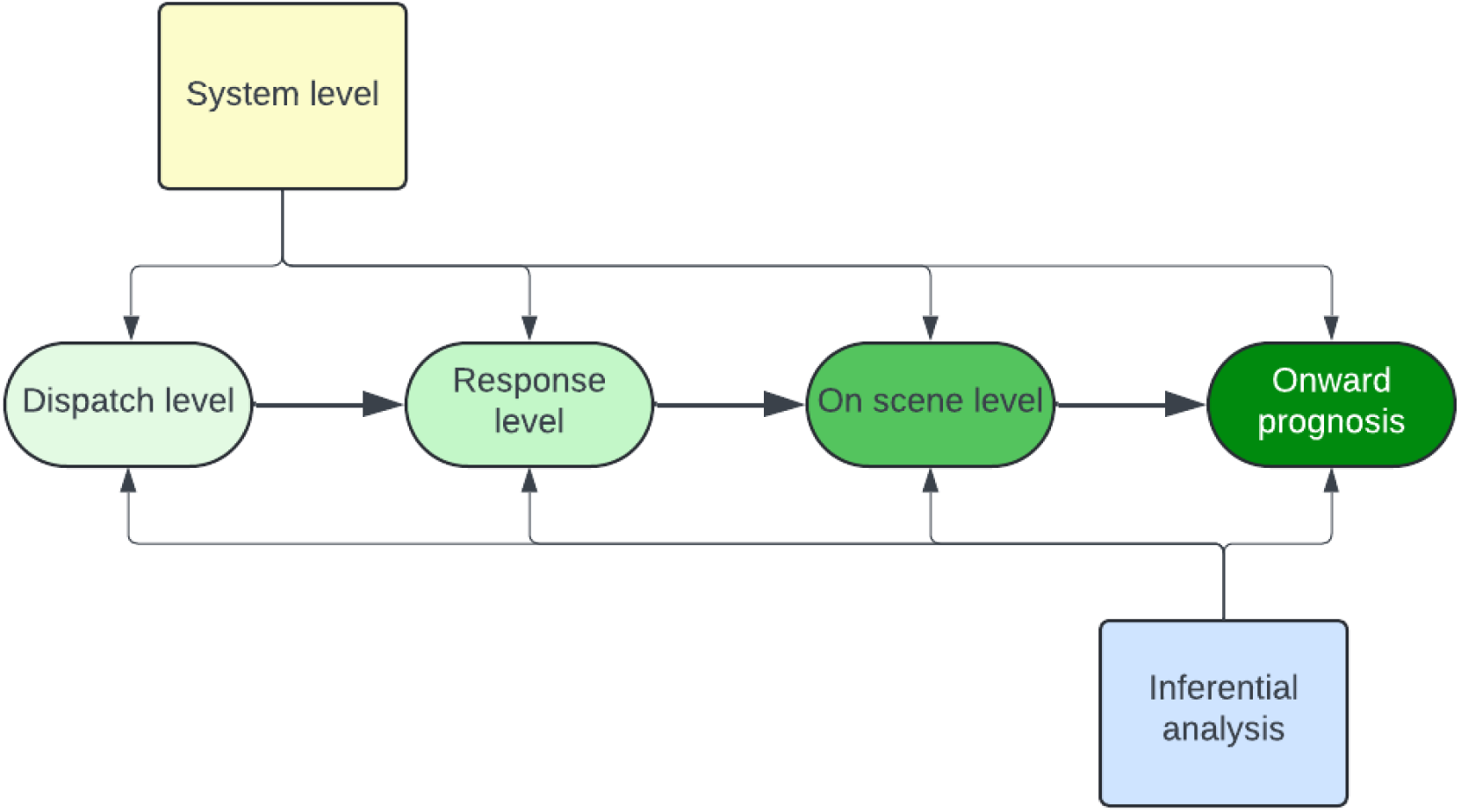

**Table 1:** A matrix that shows the count of studies clustered by applied area vs methodological type.

| Count of Level | Method Group |  |  |  |  |  |
| --- | --- | --- | --- | --- | --- | --- |
| Applied area | 1. Classical Machine Learning | 2. Deep Learning | 3. Evolutionary | 4. Natural Language | 5. Ensemble / | Grand Total |

|  |  |  |  | e | Proprietary |  |
| --- | --- | --- | --- | --- | --- | --- |
| A. System level | 14 | 17 | 11 | 0 | 4 | 46 |
| B. Dispatch Zone | 5 | 3 | 13 | 2 | 9 | 32 |
| C. Response Zone | 10 | 21 | 8 | 0 | 4 | 43 |
| D. On Scene Zone | 35 | 21 | 3 | 3 | 13 | 75 |
| E. Onward prognosis | 9 | 4 | 0 | 0 | 6 | 19 |
| F. Inferential analysis | 8 | 6 | 0 | 6 | 1 | 21 |
| **Grand Total** | **81** | **72** | **35** | **11** | **37** | **236** |

## The System Level

46 studies focused on larger-scale, macro-level applications of AI in emergency medical services (EMS). “System-level AI” refers to methods designed to optimise the whole system infrastructure across regions or jurisdictions, focusing on aggregate-scale decisions rather than individual incidents [12, 13]. Key components include forecasting, location allocation, scheduling, and resource management - requiring machine learning, optimisation, simulation, or hybrid algorithms to handle complexity and uncertainty [14].

### Use Cases of AI at the System Level

#### Demand Forecasting

For out-of-hospital services, anticipating demand and positioning resources accordingly is crucial. Studies develop predictive models to forecast EMS demand over time and space [15–18]. Others focus on specific purposes: Martin et al. [19] and Ramgopal et al. [20] employ techniques to anticipate demand surges, Lin et al. [21] explore deep learning for short-term call volume forecasting, and Kang et al. [22] predict critical care needs.^10–13^

#### EMS Resource and Station Location

After understanding demand patterns, the strategic focus shifts to balancing geographically positioned resources. Studies tackle location-allocation problems, aiming to optimise static resources (stations, defibrillator sites) and dynamic resources (ambulances, drones) to maximise coverage or minimise response times.

Approaches include:

- Integer Linear Programs and Mixed-Integer Models [23].
- Genetic Algorithms [24–27], Particle Swarm Optimisation [28], and artificial immune systems [29].
- Multi-Objective Approaches balancing cost, coverage, and response times [30–32].

#### Deployment and Routing Beyond Individual Dispatch

Several studies cover strategic routing for uncertain demand and geographical constraints, including predictive route planning for drones or autonomous ambulances and traffic/scheduling constraints [25, 27, 33, 34].

#### Integration of Climate and Environmental Factors

Some research incorporates external phenomena like heatwaves and air quality into forecasting EMS calls [15, 35], recognising that system-level modelling must account for environmental triggers alongside demographics, health trends, and temporal patterns [20, 30].

### Future Directions

#### Real-Time Adaptive Systems

Moving forward, emphasis will likely shift to real-time, adaptive models utilising traditional and emerging data sources. Continuous learning approaches could stress-test network design changes without disrupting operations [32, 36].

#### Climate Resilience and Disaster Preparedness

Climate variables can help prepare EMS systems for surges [15, 35]. Future research may combine climate modelling with epidemiological data to forecast unusual demand patterns [30, 37]. Critical incident management remains important to emergency management leaders [16].

#### Collaboration with Other Public Services

Some solutions advocate cross-agency coordination, linking EMS with firefighting, police, and public transport data [38,39]. Integrated frameworks could orchestrate multi-service dispatch in major emergencies [21, 40].

#### Equity and Ethical AI

Research increasingly highlights equitable resource allocation - Luvaanjalba & Wu [24] address rural-urban equity balance, while Chen et al. [41] consider survival challenges in remote regions. However, limited evidence exists of models including other health determinants at this strategic level. Algorithmic fairness remains challenging, with calls for open benchmarking, bias audits, and stakeholder engagement [42, 43]

### The Dispatch Zone

There were 32 studies applicable to the dispatch zone, focusing on how artificial intelligence (AI) enhances decision-making and resource allocation during emergency medical dispatch.

Dispatch-level AI typically optimises two interlinked processes: (1) Call handling—the classification and triage of emergency calls, and (2) Resource deployment—the assignment of ambulances or other emergency resources to patients; balancing coverage, response times and crew workloads [44, 45]. AI-driven dispatch systems could enhance decision-making by integrating real-time data, historical call volumes, patient acuity predictions, and geospatial information, potentially leading to faster, more precise dispatch decisions and optimised route planning [46].

### Use Cases of AI in Dispatch

#### Triage and Call Classification

Several studies explore machine learning (ML) or natural language processing (NLP) for predicting call severity and prioritisation. Chin et al. [47] discuss early recognition of emotional cues in out-of-hospital cardiac arrest calls, enabling prioritisation of emotionally unstable callers. Del Campo-Ávila et al. [48] proposed data mining techniques that aided telephone operators in detecting suicidal behaviour.

#### Ambulance Location and Deployment Optimisation

A major focus is optimal ambulance placement and relocation, anticipating demand surges. Babaei and Shahanaghi [49] adopt predictive models for demand forecasting and dynamic redeployment to minimise response times.

Mathematical models combined with heuristic algorithms support decision-making. Ravandi et al. [44] propose a multi-objective formulation considering ambulance allocation and technician capability. McCormack and Coates [50] present simulation models demonstrating increased OHCA survival rates without additional resources. Hussein, Frikha and Rahebi [51] employ optimisation algorithms showing superiority over other methods in busy urban centres.

#### Predictive Routing and Real-Time Navigation

Studies address routing challenges incorporating real-time traffic data. Talebi, Shaabani and Rabbani [52] developed a bi-objective routing model accounting for travel time and patient-critical factors. Hybrid optimisation approaches aim to decrease response time, improving patient outcomes [53].

### Accuracy and Success of Dispatch-Level AI

#### Performance Metrics

Evaluating success varies between development and real-life implementation. Research models evaluate success using accuracy, sensitivity, specificity, or area under the curve [48, 54]. Chin et al. [47] report 80-90% accuracy in distinguishing cardiac arrest calls from less urgent events.

In real-life settings, systems evaluate success via response times and coverage ratios [46]. Dynamic redeployment studies often achieve 10-20% reductions in mean response time relative to static placement. Multi-objective models demonstrate better coverage with fewer ambulances [44].

### Future Directions

#### Integration with On-Scene and Onward Prognosis Systems

Growing interest exists in linking dispatch systems with on-scene and prognosis tools for end-to-end AI-driven emergency care. Spina et al. [55] tested a model detecting COVID-19 at call triage, incorporating physiological factors for appropriate hospital referral, showing a superior performance by integrating on-scene with triage data.

#### Ethical and Equity Considerations

Concerns around fairness arise if AI dispatch systems systematically favour certain regions or inadvertently discriminate. Future work should emphasise transparency, stakeholder engagement and rigorous bias audits [56, 57].

#### Adaptive and Real-Time Updates

Future work should explore how AI can use real-time information to respond dynamically to demand surges [58]. Rautenstrauß & Schiffer [59] developed a model creating heatmaps according to ambulance demand, outperforming prior metrics by more than 9% when applied to real data.

### The Response Zone

There were 43 studies in the response zone focusing on real-time or near-real-time interventions and decision-making within the emergency response process. These approaches typically aim to streamline en-route care, pre-arrival scheduling, and dynamic control systems—serving as a bridge between initial dispatch decisions and on-scene interventions.

#### Use Cases of AI in Response

Three primary technological clusters emerge from the literature. First, smart traffic management systems employ AI to create “green corridors” for ambulances. Notable implementations include convolutional neural networks for vehicle density estimation [60], acoustic-based emergency vehicle detection with 96% accuracy [61] and fuzzy logic controllers for siren recognition with 99% effectiveness [62]. These innovations collectively reduce intersection delays by approximately 20% for priority vehicles.

Second, route optimisation algorithms address the complex problem of ambulance navigation in congested urban environments. Advanced approaches include memetic algorithms for patient transport scheduling [63], Tabu Search-based hyper-heuristics demonstrating up to 83% improvement in coverage rates [64], and swarm-intelligence methods for complex terrain navigation [65]. These algorithms adapt to changing conditions dynamically, improving dispatch efficiency and arrival time prediction.

Third, alternative response mechanisms, particularly drone-based solutions, show promising applications for out-of-hospital cardiac arrest. Mackle et al. [66] demonstrated that strategically positioned aerial ambulance drones could halve critical response times, while Boutilier and Chan [67] reported up to 76% coverage improvement in certain scenarios.

Emerging 5G connectivity solutions enhance these alternative responses through improved data throughput for telemedicine applications [68].

Collectively, these innovations represent a paradigm shift in emergency medical services, moving from reactive response to proactive optimisation. While most studies report significant performance improvements in controlled environments, future research should focus on real-world implementation challenges, including regulatory barriers and systems integration across multiple agencies and jurisdictions.

#### Future Directions

Although current findings are promising, some authors emphasise that larger-scale field studies are needed to establish external generalisability across diverse geographical contexts [66, 69]. Ftaimi & Mazri (2020) [70] Sangeetha et al. [71] and underscore the importance of broader data collection frameworks capable of handling heterogeneous traffic environments, while Halaly & Tsur [72] suggest harnessing deeper real-time learning paradigms—such as reinforcement learning—to manage dynamic conditions more effectively. Aringhieri et al. [57] also call attention to ethical and fairness considerations, emphasising that future interventions should not exacerbate existing disparities in emergency access. The practical challenges of scaling these solutions, including cost and public acceptance, are highlighted by numerous authors, indicating that implementation logistics must be carefully coordinated [67, 68, 73]. Ultimately, forging stronger collaboration among engineers, healthcare providers, and municipal authorities, as advocated by Zhang et al. [63] will be vital for transforming these algorithmic innovations into reliable, large-scale emergency response systems.

### The On Scene Zone

AI has demonstrated remarkable capabilities for predicting and classifying diagnoses whilst providing real-time recommendations to clinicians [74] during out-of-hospital care. There were 75 studies in the “on-scene zone” category involving immediate decision support for interpreting patient vitals, supporting resuscitation choices, and handling urgent transport decisions.

Machine learning, deep learning, language processing, and clinical observation interpretation aim to support clinicians’ decisions in community settings, often without specialist staff, equipment, and investigations. Modern health systems increasingly require out-of-hospital clinicians to decide whether to discharge on scene, refer onward, or transport patients to various specialist or generalist facilities [75]. AI-supported clinical decisions can potentially improve outcomes by enhancing diagnostic accuracy, optimising treatment selection, and reducing medical errors [76].

#### Use Cases of AI at the On-Scene Level Cardiac Arrest Recognition and Intervention

Several studies concentrate on out-of-hospital cardiac arrest (OHCA) scenarios, predicting outcomes such as the return of spontaneous circulation (ROSC), ECG-based pulse detection, and overall survival. Figuera et al. [77] and Isasi et al. [78] apply machine learning to differentiate shockable from non-shockable rhythms, potentially accelerating defibrillation time. Holmström et al. [79] and Hassan et al. [80] explore models alerting clinicians whether extended CPR is likely to succeed. Most studies rely on retrospective data, not accounting for real-time variables in resuscitation scenes.

#### ECG Interpretation and Acute Coronary Syndromes

Early identification of acute coronary syndrome (ACS) is imperative for intervention and specialist transport. De Koning et al. [81] propose an algorithm processing ECG signals with higher specificity than existing models. Studies explore diagnostic performance and admission prediction by combining demographic data, symptoms, and ECG waveforms [17, 82, 83]. However, research gaps remain for conditions without ECG changes and how technologies interact with clinician experience of atypical presentations [84].

#### Triage and Immediate Diagnostic Support

Some systems provide advanced triage or diagnostic prompts. Shirakawa et al. [83] developed a hospital admission prediction model using sex, chief complaint, vital signs, and medical history. Hassan et al. [80] developed algorithms suggesting appropriate facilities based on geo-mapping navigation and stroke-related timelines.

#### Accuracy and Success of On-Scene AI

Determining accuracy depends on the use case, health system context, clinician acumen, and the interplay between decision support and decision-making. Kovari [85] highlights that sector-specific evaluation requires stakeholder engagement, design process, usability, trustworthiness, and clinical validation.

Studies report sensitivity/specificity, accuracy, and AUC for condition classification. Figuera et al. [77] noted AUC values exceeding 0.90 for shockable rhythm detection, while Bouzid et al. [82] claim sensitivities around 85-90% for ECG-based ACS diagnosis. Shirakawa et al [83] and Majouni et al. [86] reference 80-90% accuracy for admission prediction.

The studies inadequately address bias, diversity, and equity considerations in defining success metrics, particularly regarding patient outcomes from underrepresented backgrounds.

#### Ethical Considerations

The most significant ethical considerations are informed consent and black-box technology. AI-augmented on-scene support raises transparency, liability, and informed consent issues. Park [87] found that disclosure of AI use in diagnosis during informed consent is generally supported; however, Xu and Shuttleworth [88] note that with “black box” algorithms, truly informed consent becomes problematic. Distinguishing AI-driven from experience-based clinical decisions further complicates matters.

These studies collectively demonstrate that on-scene AI offers potentially lifesaving advantages while identifying research gaps in real-time testing, understanding of atypical presentations, and considerations of equality, diversity, and informed consent in patient care.

### The Onward Prognosis

Nineteen studies examined onward prognosis, forecasting future patient states based on out-of-hospital or early clinical data.’Onward prognosis’ denotes the application of artificial intelligence (AI) methodologies to predict a patient’s likely clinical course. These studies address outcomes including survival, neurological recovery and hospital admission, reducing many predictors for OHCA outcomes and reducing bias [89–92]. Rather than merely classifying a patient’s present state, onward prognosis provides actionable insights into condition evolution, enabling better resource allocation and informed care decisions [93].

### Use Cases of AI for Onward Prognosis

#### Out-of-Hospital Cardiac Arrest (OHCA) Survival Prediction

Many studies target OHCA, forecasting survival to hospital discharge or neurologically intact survival. Classical machine learning models predict outcomes by leveraging patient demographics, resuscitation times and clinical indicators [94, 95]. Several studies adopt similar approaches, highlighting novel feature sets or ensemble techniques (89-93, 96-102]. These models guide clinicians in understanding which patients may benefit from prolonged resuscitation efforts.

Beyond OHCA survival, studies identify neurological intact survival as a separate or secondary endpoint [90, 91, 93, 98,100].

#### Short-Term Mortality and Neurological Outcomes

Two studies use machine learning models to focus on general short-term mortality predictions for critically ill patients [102, 103]. Venema et al. [104] examined prediction outcomes for triaging out-of-hospital stroke care strategies supporting specialist centre transfers, similar to Moon et al. [99] for OHCA interventions.

#### Hospital Admission Forecasting

A smaller subset addresses whether patients require hospital admission or can be safely discharged. Strum et al. [105] developed an ensemble model for forecasting hospital admission from out-of-hospital data to streamline patient flow and reduce overcrowding. Real-time prediction allows EMS systems to optimise facility selection and initiate advanced notifications to in-hospital teams.

#### Accuracy and Success of Predictive Models Reported Performance Metrics

The studies employ various modelling techniques—classical machine learning [89, 94–97, 103, 104, 106, 107], deep learning [98–101] and ensemble methods [90–93, 102]. Evaluation metrics include area under the receiver operating characteristic curve (AUC), sensitivity, specificity, and positive predictive value (PPV).

Multiple OHCA survival studies achieve AUC values in the 0.80-0.90 range [90, 91, 93, 94, 98, 101, 107].

For short-term mortality, Tamminen et al. [103] indicate high discriminative ability (AUC above 0.85) using vital signs and demographics. Regarding neurological outcomes, Kwon et al. [98] demonstrate that deep-learning models can effectively classify recovery categories with sensitivity and specificity above 80%.

### Future Directions

#### Data Integration and Real-Time Deployment

Greater emphasis on real-time data integration from multiple sources (wearable devices, ambulance telemetry, electronic health records) could significantly improve prediction timeliness and accuracy [102]. Studies suggest focusing on specific, reliable predictors to support real-time data integration [92, 93]. Early warnings triggered en route would allow specialised equipment or teams to be prepared before patient arrival [99, 104, 105].

#### Generalisability and Transparency

Future work should improve the interpretability of AI-driven prognosis tools and validate them across diverse populations. Model transparency is essential for building trust among EMS personnel and regulatory bodies [92, 105]. For generalisability, better calibration and understanding of real-time machine learning applications [92].

#### Beyond Mortality to Holistic Outcomes

There is a recognised need to expand beyond binary survival metrics to incorporate patients’ functional status, quality of life, and social determinants of health, ensuring comprehensive outcome assessment [99, 104].

### The Inferential Analysis

There were 21 studies at the inferential analysis level, providing higher-level insights through secondary analyses of prehospital data. These studies investigate trends, associations, and underlying mechanisms—exploring social, demographic, and health system factors influencing EMS.

Inferential analysis examines how variables correlate within EMS datasets [108, 109], seeking to understand why certain outcomes occur and gleaning insights into disparities in prehospital care [110], climate impacts on call volumes [111] and how scene times relate to patient outcomes [112].

#### Use Cases of AI for Inferential Analysis Social Determinants of Health and Disparities

Studies reveal inequities affecting EMS usage. Monlezun et al. [110] identify differential OHCA survival rates among populations, suggesting targeted efforts could improve outcomes. Using NLP, Tignanelli et al. [113] and Burnett et al. [114] explore neighbourhood factors influencing prehospital intervention quality.

#### Public Health and Population-Level Phenomena

EMS data can examine broader public health issues. Ajumobi et al. [115] and Graham et al. [116] identify opioid overdoses through paramedic records for public health surveillance. Mohammadi et al. [111] simulate how climate factors might elevate call demand.

#### Scene Times and Protocol Efficacy

Studies investigate how prehospital timescales affect outcomes. Shin et al. [112] link on-scene intervals with survival outcomes. Kawai et al. [117] explore transport time effects on resuscitation after OHCA. Harmon et al. [118] and Choi et al. [119] note that machine learning outperforms rule-based methods for specific condition identification.

#### Natural Language Processing (NLP) and Data Extraction

Redfield et al. [120] link EMS and ED records to extract clinical factors. Tohira et al. [121] and Tignanelli et al. [113] classify patient sub-cohorts to identify prevention opportunities. Silverman et al. [122] identify interventions correlating with favourable outcomes.

#### Key Findings and Future Directions

Multiple works confirm socioeconomic disparities in response times and survival [110, 114]. Some studies reveal that appropriate scene time management improves neurological outcomes [112, 121].

Future directions include integrating social and environmental data with EMS logs [112], enhancing causal inference methodologies [109] and developing ethical frameworks for data usage [123]. Translating insights into operational changes requires collaboration between data scientists, clinicians, and EMS leadership [124].

These inferential contributions explore underlying drivers in EMS data, offering valuable perspectives on public health, clinical outcomes, and system equity.

## Discussion

### Summary of evidence

This scoping review identified 236 publications describing potential AI applications across five domains of out-of-hospital care: system level (46 studies), dispatch (32 studies), response (43 studies), on-scene (75 studies), and onward prognosis (19 studies); with further inferential applications also. While the literature reveals numerous theoretical use cases spanning the entire pathway, a striking finding is a substantial gap between proposed applications and actual implementation in real-time clinical practice.

System-level applications predominantly focus on demand forecasting and resource optimisation, with theoretical coverage improvements of 10-20% reported in simulation studies compared to standard approaches. Similarly, dispatch-level AI models project response time reductions of up to 10-20%, and response zone applications suggest travel time reductions of 15-30% in controlled environments. On-scene applications demonstrate promising accuracy in retrospective analyses, with cardiac arrest rhythm detection achieving AUC values exceeding 0.90 and ACS prediction sensitivities of 85-90%. In validation datasets, onward prognosis models predict patient outcomes with AUC values of 0.80-0.90.

However, our review found that only a small fraction of these promising applications have progressed beyond conceptual models, simulations, or retrospective analyses to real-world implementation. Of the 236 studies reviewed, fewer than 15% reported functional deployments in active clinical settings, and even fewer (<5%) documented sustained implementation with longitudinal evaluation. This implementation gap represents one of the most significant barriers to realising the potential benefits of AI in settings.

### The implementation gap: barriers to real-world application

A critical finding from our review is the substantial gap between AI development and practical implementation in settings. Despite the abundance of promising research, few solutions have successfully transitioned from laboratory to clinical practice. This implementation gap appears to stem from several key barriers:

1. **Technical challenges**: Many studies highlight difficulties in data integration across disparate systems, with incompatible formats and interoperability issues preventing seamless information flow. For example, Blomberg et al. [125] noted that promising AI tools remained unused due to integration challenges with existing clinical systems.
2. **Regulatory and governance hurdles**: The regulatory landscape for AI in healthcare remains uncertain, with few studies addressing compliance with medical device regulations or data protection frameworks. This regulatory ambiguity creates hesitancy among healthcare providers to adopt novel technologies.
3. **Validation and evidence requirements**: Healthcare systems typically require robust evidence of clinical and economic benefits before adopting new technologies. Most studies in our review provided limited evidence of real-world effectiveness, focusing instead on technical performance metrics in controlled settings.
4. **Organisational barriers**: Successful implementation requires substantial organisational change, including workflow redesign, staff training, and leadership support. Few studies addressed these critical change management aspects of technology implementation.

The rare examples of successful implementation, such as Spina et al. [55], who integrated call triage data with on-scene physiological measurements to improve COVID-19 patient routing, highlight the importance of addressing these barriers systematically and collaboratively.

### Ethical considerations and implementation challenges

Our review identified recurring ethical considerations across domains, particularly regarding algorithmic transparency, equity, and informed consent. Many advanced AI models’ are “black box” in nature, especially in on-scene clinical decision support, which raises important questions about accountability and trust. Hassan et al. [80] and Park [87] emphasise the importance of transparent decision-making processes, particularly when AI influences life-critical interventions.

Equity concerns are evident across all domains, with studies by Monlezun et al. [110] and Burnett et al. [114] revealing how AI systems may inadvertently perpetuate or exacerbate existing disparities in emergency care access and outcomes. Few studies explicitly address how AI implementations might affect underserved populations or regions with limited technological infrastructure.

Implementation challenges highlighted across studies include data quality and integration issues, resistance to adoption among clinical staff, and the need for robust validation in diverse real-world settings. Blomberg et al. [125] noted low utilisation rates of decision support tools despite clinician acknowledgement of their utility, suggesting that technological capability alone is insufficient for successful implementation.

### Limitations

This review has several limitations. First, our search strategy may not have captured all relevant studies, particularly those published in languages other than English or specialised technical journals. Second, the rapid evolution of AI technologies means that more recent innovations may not yet be represented in the published literature. Third, the heterogeneity of study designs, outcome measures, and reporting standards made direct comparisons challenging, particularly regarding clinical impact.

Furthermore, publication bias likely favours positive results, potentially overestimating the benefits of AI technologies in settings. Many studies report algorithm performance in controlled or retrospective scenarios, which may not translate to real-world effectiveness. Finally, our categorisation of studies into five domains, while useful for organising findings, may oversimplify the complex interrelationships between different aspects of the pathway.

## Conclusions

This scoping review reveals a paradox in the field of AI for out-of-hospital care: an abundance of theoretical applications with promising performance metrics, but a scarcity of successful real-world implementations. While the literature demonstrates significant potential for AI across the pathway, the field faces a critical implementation gap that must be addressed to realise these benefits in clinical practice.

Future research and implementation efforts should focus on:

1. **Bridging the implementation gap** by creating implementation science frameworks specifically for AI in emergency care, with clear pathways from development to deployment and evaluation.
2. **Building evidence for clinical impact** beyond technical performance metrics with pragmatic trials that assess patient outcomes, staff experience, and system-level effects in real-world settings.
3. **Addressing interoperability challenges** through standardised data formats and integration protocols that enable information flow across the pathway.
4. **Developing implementation toolkits** that address organisational, regulatory, and technical barriers to adoption, including change management strategies and stakeholder engagement approaches.
5. **Creating collaborative implementation networks** that bring together technology developers, clinical teams, patients, and implementation scientists to tackle the complex challenges of moving from proof-of-concept to sustained clinical use.

By shifting focus from developing novel AI applications to implementing and evaluating existing ones in clinical practice, the emergency care community can begin to close the implementation gap and harness the transformative potential of these technologies to improve patient care. Without this shift, the field risks accumulating an ever-growing library of promising but unused AI solutions while emergency care systems continue to face mounting pressures.

## Data Availability

All data produced in the present work are contained in the manuscript

