## Supplementary Material for "The use of Artificial Intelligence in the out of hospital care settings: A Scoping Review"

**Supplementary Material: Detailed Methods**

**Complete Search Strategy**

The complete search strategy included the following terms:

**Out-of-Hospital Care Terms:**

"Emergency Medical Services"[MeSH] OR "Emergency Medical Services" OR EMS OR "Prehospital Care" OR "Ambulances"[MeSH] OR Ambulance* OR Paramedic* OR "First Responder*" OR "Emergency Treatment"[MeSH] OR "Out-of-Hospital"

**Artificial Intelligence Terms:**

"Artificial Intelligence"[MeSH] OR "Artificial Intelligence" OR AI OR "Machine Learning"[MeSH] OR "Machine Learning" OR "Deep Learning"[MeSH] OR "Deep Learning" OR "Neural Networks (Computer)"[MeSH] OR "Neural Network*" OR "Natural Language Processing"[MeSH] OR "Natural Language Processing" OR NLP OR "Predictive Analytics" OR "Data Mining"[MeSH] OR "Data Mining" OR Algorithm* OR "Intelligent Systems"

The search strategy was adapted for each database while maintaining the core concept structure. Date limits were set from January 1, 2013, to March 31, 2024.

**Detailed Data Extraction Template**

The data extraction template included the following fields:

**Study Identification**

- Author(s)
- Year of publication
- Country of origin
- Journal/Conference/Preprint server

**Study Characteristics**

- Study design
- Sample size (if applicable)
- Study duration
- Funding source

**AI Technology Details**

- Type of AI/ML approach (e.g., supervised learning, deep learning, natural language processing)
- Algorithm(s) used
- Training dataset characteristics
- Validation approach

**Emergency/Urgent Care Context**

- Type of service (ambulance, NHS 111, etc.)
- Healthcare system context
- Target population
- Healthcare setting specifics

**Implementation Details**

- Implementation stage (conceptual, prototype, pilot, fully implemented)
- Integration with existing systems
- User interface/experience considerations
- Implementation barriers and facilitators

**Outcome Measurements**

- Primary outcomes
- Secondary outcomes
- Evaluation metrics
- Comparison/control (if applicable)

**Results and Findings**

- Key quantitative results
- Key qualitative findings
- Author-reported limitations
- Author-reported implications

**Network Analysis Methodology**

Network analysis of author collaborations was conducted using VOSviewer (v1.6.18). A co-authorship network was constructed where nodes represented individual authors and edges represented collaboration links between authors.

We calculated the following metrics:

- Network density: The proportion of potential connections that are actual connections
- Degree centrality: The number of direct connections each author has
- Betweenness centrality: The extent to which an author lies on paths between other authors

Clusters were identified using the default clustering algorithm in VOSviewer with a resolution parameter of 1.0. Visualization was generated with nodes sized according to the number of publications and colored according to cluster membership.
